# Developmental trajectories in infants and pre-school children with Neurofibromatosis 1

**DOI:** 10.1101/2023.12.05.23299174

**Authors:** Hannah Slevin, Fiona Kehinde, Jannath Begum-Ali, Ceri Ellis, Emma Burkitt-Wright, Jonathan Green, Mark H. Johnson, Greg Pasco, Tony Charman, Emily J.H Jones, Shruti Garg, the EDEN-STAARS team

## Abstract

**Objective:** This prospective cohort study examines the cognitive, behavioural, ADHD trait and autism symptom development in infant and pre-school children with Neurofibromatosis 1 (NF1) compared with typically developing (TD) children without a family history of neurodevelopmental conditions.

**Methods:** Data from standardised tests was gathered at 5, 10, 14, 24 and 36 months of age (NF1 n=35, TD n=29). Developmental trajectories of cognitive and adaptive behavioural development from 5 to 36 months were analysed using linear mixed modelling. Measures of ADHD and autism traits were assessed at 24 and 36 months.

**Results:** The developmental trajectory of cognitive skills (all domains of the Mullen Scales of Early Learning -MSEL) and behavioural skills (four domains of the Vineland Adaptive Behaviour Scale -VABS) differed significantly between NF1 and TD groups. Post-hoc tests demonstrated that the NF1 participants scored significantly lower than TD participants at 24 months on all MSEL and VABS domains. The NF1 cohort demonstrated higher mean autism and ADHD traits at 24 months and 14% of the NF1 cohort met a research diagnostic classification for autism at 36 months.

**Conclusion:** By 24 months of age, the NF1 cohort show lower cognitive skills and adaptive behaviour and higher levels of autism and ADHD traits as compared to TD children.

## INTRODUCTION

Neurofibromatosis 1 (NF1) is a single gene neurocutaneous condition, with a birth incidence of 1 in 2700.^1^ 50% of cases arise through autosomal dominant inheritance from a parent, and 50% via a sporadic pathogenic variant of the tumour-suppressor NF1 gene on chromosome 17q11.2. This gene codes for the protein neurofibromin, which plays a role in regulating neuronal cell development.^2^ NF1 is characterised by phenotypic variability, and clinical diagnosis is based on the revised National Institute of Health (NIH) criteria.^3^

Many children with NF1 experience cognitive and behavioural difficulties which impact their quality of life. Overall intellectual ability is only slightly lower than children who do not have NF1, however specific cognitive deficits impact perceptual (particularly visuospatial) skills, executive functioning, adaptive behaviour, attention and motor skills.^4,5^ Research also indicates high rates of co-occurring conditions such as Autism Spectrum Disorder (henceforth referred to as autism) (29%) and Attention Deficit Hyperactivity Disorder (ADHD) (50%).^6,7^

The majority of studies analysing the cognitive, social and behavioural phenotype in NF1 have concentrated on school-aged children. Several cross-sectional studies have focused on the pre-school period, which consistently demonstrate that young children with NF1 show cognitive, behavioural and social differences compared to age-matched controls.^2,4,8-12^ However, the age and sequence at which these differences begin to emerge is not fully understood. Understanding this is crucial for identifying early markers of later neurodevelopmental outcomes, and has clinical implications for screening and intervention, particularly educational planning.^13^

Two previous studies have analysed trajectories of cognitive development in toddlers.^14,15^ Lorenzo et al (2015) assessed the early development of 39 children with NF1 aged 21, 30 and 40 months compared with matched controls; the NF1 cohort demonstrated lower cognitive functioning than controls.^14^ Wessel et al (2013) assessed 124 infants with NF1 at a range of ages (0-8) using parental report measures. Gross and fine motor delays were found to emerge aged 3-5, whereas academic delays tended to present at a later age.^15^ However, the majority of the cohort had just one assessment.

Previous papers from our group examined trajectories of cognitive and adaptive behavioural development in infants with NF1 aged 5 to 14 months compared with typically developing (TD) children.^13^ At this early stage in development, no group differences were observed in trajectories of cognitive and adaptive functioning, nor differences on social communication measures. However, early differences in neural processing including auditory processing and excitation/inhibition balance were observed, both of which were related to later autistic traits.^16,17^

Our study aim is to examine the early cognitive, behavioural, autism and ADHD trait development of infants and pre-school aged children with NF1, compared with a cohort of TD children. Our objectives were to build on the earlier work of our team^13^ by examining longitudinal trajectories of cognitive and behavioural development from 5-36 months, and ADHD trait development at 24 and 36 months. We also aimed to examine emergence of autism traits at 24 and 36 months.

We hypothesised that children with NF1 would have lower cognitive and adaptive functioning over time, and higher levels of ADHD and autism traits compared with TD children. Based on our previous work, we interrogate whether differences begin to emerge at age 24 or 36 months. To our knowledge, this is the first study to examine cognitive and behavioural development from infancy in children with NF1 using both parental report and objective assessment measures. This prospective approach is critical in avoiding ascertainment bias seen with older children, where participants may be more likely to participate if they are experiencing developmental delays.

## METHODS

The Early Development in NF1 (EDEN) study is a UK-based prospective longitudinal cohort study investigating early development in infants and children with NF1. Participants were enrolled through regional genetic centres and NF1 charities. Participants in the TD group were recruited from the Studying Autism and ADHD in at Risk Siblings (STAARS) study at the Centre for Brain and Cognitive Development, Birkbeck, University of London. Participants were assessed at 5, 10, 14, 24 and 36 months of age. Rolling recruitment was conducted between 2016-2019.

Inclusion criteria for the NF1 cohort included a) infants up to 16 months of age at the time of recruitment b) NF1 diagnosed via testing of cord blood samples or clinical diagnosis.

Inclusion criteria for the TD group included a) infants up to 16 months of age at the time of recruitment b) no first-degree relatives with known genetic conditions, autism or Attention Deficit Hyperactivity Disorder (ADHD) c) no parent-reported developmental issues d) full-term birth (gestational age at least 36 weeks).

Exclusion criteria for both groups included a) conditions which might make it difficult for the infant to participate, including serious physical complications of NF1 b) significant hearing or visual impairments c) significant prematurity d) parents with significant learning difficulties or who were unable to give informed consent.

Procedural details are included in **Supplementary materials**.

### Measures

#### Maternal education

Maternal education was classified as either primary, secondary, undergraduate or postgraduate (1,2,3 or 4) (**Table 1**). This has been shown to be significantly associated with trajectories of cognitive and adaptive functioning at 5, 10 and 14 months in this population.^15^

**Table 1:** Demographic characteristics and number of participants at each Timepoint

|  | 5 Months |  |  |  | 10 Months |  |  |  | 14 Months |  |  |  | 24 months |  |  |  | 36 months |  |  |  |
| --- | --- | --- | --- | --- | --- | --- | --- | --- | --- | --- | --- | --- | --- | --- | --- | --- | --- | --- | --- | --- |
|  | TD group |  | NF1 Group |  | TD group |  | NF1 Group |  | TD group |  | NF1 Group |  | TD group |  | NF1 Group |  | TD group |  | NF1 Group |  |
|  | *n* | *Mean*<br>*[SD]* | *n* | *Mean*<br>*[SD]* | *n* | *Mean*<br>*[SD]* | *n* | *Mean*<br>*[SD]* | *n* | *Mean*<br>*[SD]* | *n* | *Mean*<br>*[SD]* | *n* | *Mean*<br>*[SD]* | *n* | *Mean*<br>*[SD]* | *n* | *Mean*<br>*[SD]* | *n* | *Mean*<br>*[SD]* |
| Age at time of testing in days | 26 | 179.19<br>[14.05] | 15 | 194.73<br>[17.85] | 27 | 321.93<br>[16.70] | 23 | 327<br>[17.11] | 23 | 447.7<br>[18.37] | 27 | 449.74<br>[23.41] | 24 | 762.25<br>[36.07] | 29 | 795.41<br>[90.98] | 20 | 1135.45<br>[55.31] | 30 | 1255.93<br>[164.87] |
| Sex (n) M/F | 26 | 17/9 | 15 | 8/7 | 27 | 16/11 | 23 | 12/11 | 23 | 13/10 | 27 | 13/ 14 | 24 | 13/11 | 29 | 14/15 | 20 | 12/8 | 30 | 14/16 |
| Maternal education (n)<br>(Secondary/undergrad/<br>postgrad) | 23 | 2/8/13 | 12 | 8/2/2 | 26 | 2/10/14 | 23 | 15/5/3 | 23 | 2/7/14 | 25 | 14/8/3 | 23 | 2/8/13 | 26 | 15/8/3 | 19 | 1/7/11 | 26 | 16/7/3 |

#### Cognitive and adaptive behavioural skills

The Mullen Scales of Early Learning (MSEL),^18^ a standardised assessment for children aged up to 68 months, was used to assess cognitive functioning at all five time points. Five domains were assessed, including Visual Reception, Fine Motor, Receptive Language, Expressive Language skills (all measured as T-scores) and an Early Learning Composite (ELC) (Standard Score).

The Vineland Adaptive Behavior Scale (VABS),^19^ a parent-report questionnaire, was used to assess adaptive behavioural skills at all five time points. Five domains were assessed, including Communication, Daily Living Skills, Socialisation, Motor skills and an Adaptive Behaviour Composite score. Analyses used Standard Scores.

#### ADHD traits

The Child Behaviour Checklist (CBCL) for ages 1.5-5,^20^ a parent-report questionnaire, was used to assess ADHD traits (inattention/hyperactivity) at 24 and 36 month time points. T-scores were used for the DSM-orientated Attention Deficit Hyperactivity problems scale. T-scores of 70 are in the clinically significant range, and 65-69 considered borderline.

#### Autism traits

The Autism Diagnostic Observation Schedule (ADOS-2), a semi-structured assessment of social communication, social interaction and imaginative play for individuals suspected to have autism,^21,22^ was utilised at 24 and 36 months. Due to social distancing constraints necessitated by the Covid-19 pandemic, some assessments instead utilised the Brief Observation of Symptoms of Autism for Minimally Verbal children (BOSA-MV), an observational measure designed to be administered remotely.^23^ **Table 3** outlines the participant numbers at each timepoint who were administered ADOS-2 versus BOSA.

**Table 2:**
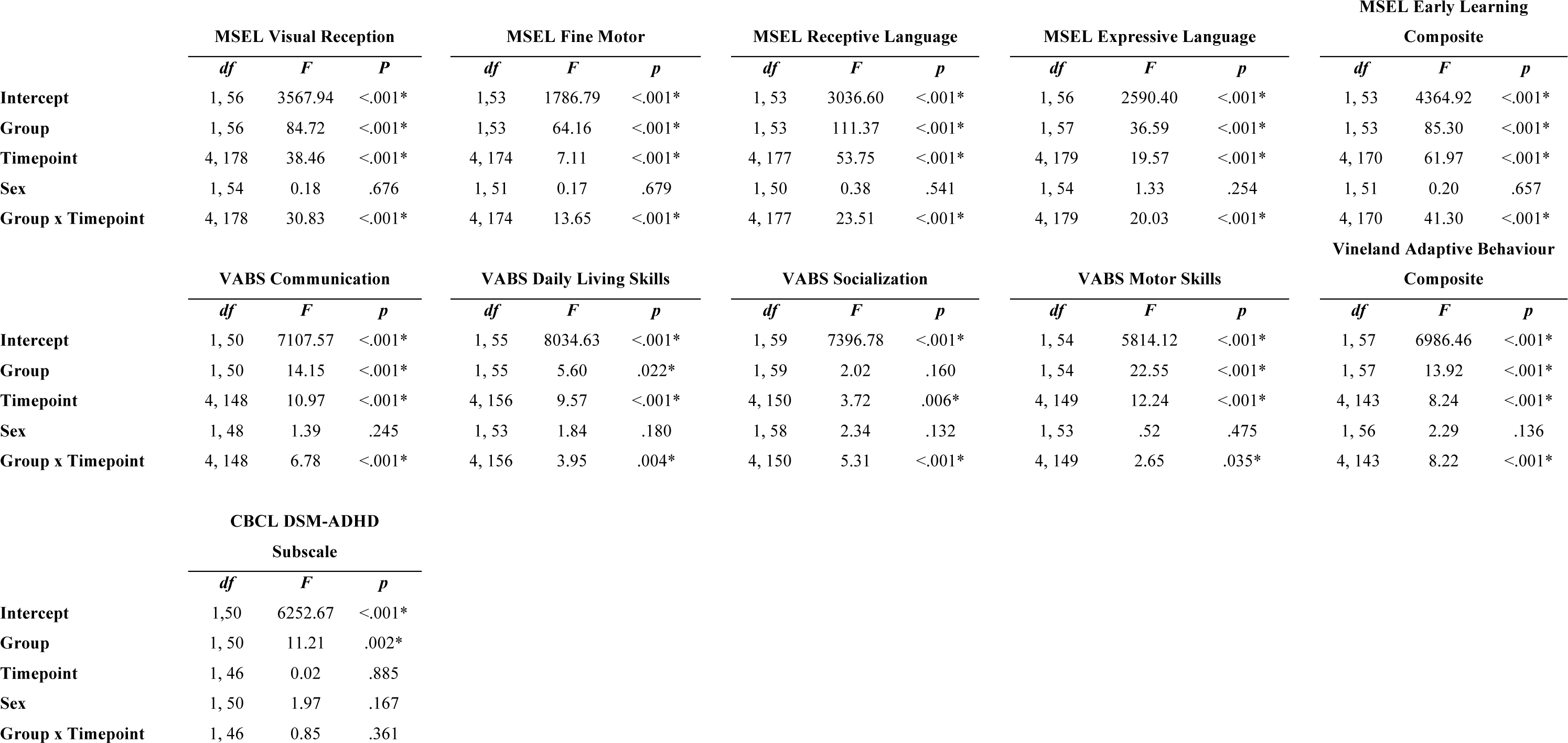
Linear mixed modelling F statistic and p values of fixed effects for all the outcome measures. Maternal education was included in all models as a covariate. df= degrees of freedom, F statistic, p value, *= significant results

|  | MSEL Visual Reception |  |  | MSEL Fine Motor |  |  | MSEL Receptive Language |  |  | MSEL Expressive Language |  |  | MSEL Early Learning Composite |  |  |
| --- | --- | --- | --- | --- | --- | --- | --- | --- | --- | --- | --- | --- | --- | --- | --- |
|  | *df* | *F* | *P* | *df* | *F* | *p* | *df* | *F* | *p* | *df* | *F* | *p* | *df* | *F* | *p* |
| **Intercept** | 1, 56 | 3567.94 | <.001* | 1,53 | 1786.79 | <.001* | 1, 53 | 3036.60 | <.001* | 1, 56 | 2590.40 | <.001* | 1, 53 | 4364.92 | <.001* |
| **Group** | 1, 56 | 84.72 | <.001* | 1,53 | 64.16 | <.001* | 1, 53 | 111.37 | <.001* | 1, 57 | 36.59 | <.001* | 1, 53 | 85.30 | <.001* |
| **Timepoint** | 4, 178 | 38.46 | <.001* | 4, 174 | 7.11 | <.001* | 4, 177 | 53.75 | <.001* | 4, 179 | 19.57 | <.001* | 4, 170 | 61.97 | <.001* |
| **Sex** | 1, 54 | 0.18 | .676 | 1, 51 | 0.17 | .679 | 1, 50 | 0.38 | .541 | 1, 54 | 1.33 | .254 | 1, 51 | 0.20 | .657 |
| **Group x Timepoint** | 4, 178 | 30.83 | <.001* | 4, 174 | 13.65 | <.001* | 4, 177 | 23.51 | <.001* | 4, 179 | 20.03 | <.001* | 4, 170 | 41.30 | <.001* |
|  | VABS Communication |  |  | VABS Daily Living Skills |  |  | VABS Socialization |  |  | VABS Motor Skills |  |  | Vineland Adaptive Behaviour Composite |  |  |
|  | *df* | *F* | *p* | *df* | *F* | *p* | *df* | *F* | *p* | *df* | *F* | *p* | *df* | *F* | *p* |
| **Intercept** | 1, 50 | 7107.57 | <.001* | 1, 55 | 8034.63 | <.001* | 1, 59 | 7396.78 | <.001* | 1, 54 | 5814.12 | <.001* | 1, 57 | 6986.46 | <.001* |
| **Group** | 1, 50 | 14.15 | <.001* | 1, 55 | 5.60 | .022* | 1, 59 | 2.02 | .160 | 1, 54 | 22.55 | <.001* | 1, 57 | 13.92 | <.001* |
| **Timepoint** | 4, 148 | 10.97 | <.001* | 4, 156 | 9.57 | <.001* | 4, 150 | 3.72 | .006* | 4, 149 | 12.24 | <.001* | 4, 143 | 8.24 | <.001* |
| **Sex** | 1, 48 | 1.39 | .245 | 1, 53 | 1.84 | .180 | 1, 58 | 2.34 | .132 | 1, 53 | .52 | .475 | 1, 56 | 2.29 | .136 |
| **Group x Timepoint** | 4, 148 | 6.78 | <.001* | 4, 156 | 3.95 | .004* | 4, 150 | 5.31 | <.001* | 4, 149 | 2.65 | .035* | 4, 143 | 8.22 | <.001* |
|  | CBCL DSM-ADHD Subscale |  |  |  |  |  |  |  |  |  |  |  |  |  |  |
|  | *df* | *F* | *p* |  |  |  |  |  |  |  |  |  |  |  |  |
| **Intercept** | 1,50 | 6252.67 | <.001* |  |  |  |  |  |  |  |  |  |  |  |  |
| **Group** | 1, 50 | 11.21 | .002* |  |  |  |  |  |  |  |  |  |  |  |  |
| **Timepoint** | 1, 46 | 0.02 | .885 |  |  |  |  |  |  |  |  |  |  |  |  |
| **Sex** | 1, 50 | 1.97 | .167 |  |  |  |  |  |  |  |  |  |  |  |  |
| **Group x Timepoint** | 1, 46 | 0.85 | .361 |  |  |  |  |  |  |  |  |  |  |  |  |

**Table 3:** Descriptive statistics including means and SDs for the NF1 and TD group at 24 and 36 month time points for ADI-R, ADOS, BOSA and CBCL

|  | 24 months |  |  |  | 36 months |  |  |  |
| --- | --- | --- | --- | --- | --- | --- | --- | --- |
|  | TD group |  | NF1 Group |  | TD group |  | NF1 Group |  |
|  | *n* | *Mean*<br>*[SD]* | *n* | *Mean*<br>*[SD]* | *n* | *Mean*<br>*[SD]* | *n* | *Mean*<br>*[SD]* |
| **Autism Diagnostic Interview - Revised (ADI-R) Diagnostic Algorithm** |  |  |  |  |  |  |  |  |
| Qualitative Abnormalities in Reciprocal Social Interaction (A) |  |  |  |  | 18 | 0.94(1.00) | 30 | 5.23(4.79) |
| Qualitative Abnormalities in Communication (B) |  |  |  |  | 18 | 0.67 (0.97) | 30 | 5.03(4.42) |
| Restricted, Repetitive and Stereotyped Patterns of Behaviour (C) |  | N/A |  | N/A | 18 | 0.39 (0.61) | 30 | 2.13(2.60) |
| Abnormality of Development Evident at or Before 36 Months (D) |  |  |  |  | 18 | 0.06 (0.24) | 30 | 1.83(1.34) |
| Meeting threshold for autism on ADI-R |  |  |  |  | 18 | 0 (0%) | 30 | 5 (17%) |
| Not meeting threshold for autism on ADI-R |  |  |  |  | 18 | 18 (100%) | 30 | 25 (83%) |
| **Autism Diagnostic Observation Schedule-2 (ADOS-2)** |  |  |  |  |  |  |  |  |
| Module (n) (Toddler/Module 1/Module 2) | 24 | 24,0,0 | 24 | 21,3,0 | 20 | 0,0,20 | 17 | 0,16,1 |
| Total scores | 24 | 3.67(1.95) | 24 | 8.08(6.64) | 20 | 4.35(3.42) | 17 | 4.47(3.86) |
| Social Affect scores | 24 | 3.13(1.80) | 24 | 6.83(5.75) | 20 | 3.60(2.84) | 17 | 3.65(3.02) |
| Restrictive and Repetitive Behaviour scores | 24 | 0.54(0.72) | 24 | 1.25(1.29) | 20 | 0.80(0.95) | 17 | 0.82(1.24) |
| Number reaching threshold for Autism Spectrum Disorder or Autism on ADOS-2 | 24 | 0 (0%) | 24 | 11 (46%) | 20 | 5 (25%) | 17 | 4 (24%) |
| **Brief Observation of Symptoms of Autism (BOSA-MV)** |  |  |  |  |  |  |  |  |
| Module (n) (Toddler/Module 1) |  |  | 3 | 2,1 |  |  | 12 | 0,12 |
| Total scores | 0 | 0,0 | 3 | 12.33(4.93) | 0 | 0,0 | 12 | 8.17(2.66) |
| Algorithm scores |  |  | 3 | 6.00(2.00) |  |  | 12 | 4.42(1.44) |
| Number reaching threshold for Autism Spectrum Disorder on the BOSA |  |  | 3 | 2 (67%) |  |  | 12 | 5 (42%) |
| **ADOS/BOSA combined** |  |  |  |  |  |  |  |  |
| Number of participants reaching threshold for Autism | 24 | 0 (0%) | 27 | 13 (48%) | 20 | 5 (25%) | 29 | 9 (31%) |
| Number of participants not reaching threshold for Autism | 24 | 24 (100%) | 27 | 14 (52%) | 20 | 15 (75%) | 29 | 20 (69%) |
| **Number of participants reaching threshold for Autism on both ADI-R and ADOS/BOSA** |  |  |  |  |  |  |  |  |
|  |  | N/A |  | N/A | 18 | 0 (0%) | 29 | 4 (14%) |
| **Child Behaviour Checklist ADHD subscale** |  |  |  |  |  |  |  |  |
| T score | 22 | 51.86(2.49) | 27 | 55.56(7.84) | 21 | 51.10(1.45) | 28 | 57.14(6.88) |

The Autism Diagnostic Interview-Revised (ADI-R), an investigator-based semi-structured interview for parents,^24^ was carried out at 36 months.

The **Supplementary Materials** outline diagnostic thresholds on the ADOS-2, BOSA and ADI-R, and detail how a research classification of autism was determined at 36 months.

### Statistical analyses

Statistical analyses were performed using IBM SPSS Statistics 28.0.0.0. Linear mixed modelling was used to analyse the change in cognition (MSEL), adaptive behaviour (VABS) and ADHD traits (CBCL DSM-ADHD subscale) over time. For each subscale, overall group differences were modelled using fixed effects (group, timepoint and sex) and random effects (ID - individual variation). Maternal education was included as a co-variate within the model.^15^ Maternal education significantly differed between groups, with mothers of TD children more likely to have a post-graduate education (Median NF1: 2, TD: 4 X^2^ (2) = 19.79, p<0.001).

In all models, sex was non-significant (**Table 2)**. The NF1 group was significantly older than the TD group at 5 months (t = 3.09, 95% CI 5.36 to 25.72 days, p = 0.004, *d* 1.00) and at 36 months (t = 3.70, 95% CI 54.60 to 186.36 days, p = <0.001, *d* 0.91). Age in days was not included in the model as a fixed effect, as this had already been corrected for by using age-corrected T scores (MSEL and CBCL) and age-corrected standard scores (VABS). Post hoc T-tests were carried out to further explore group differences on the MSEL and VABS at each timepoint.

Missing data were imputed using the maximum likelihood option. A p value of <0.05 was considered as significant. For post-hoc tests, a corrected p value of <0.01 was considered significant. Pearson Chi-squared tests were carried out for proportion of participants meeting autism threshold on the ADOS-2/BOSA/ADI-R (as outlined in the **Supplementary materials)**. Mann-Whitney non-parametric analyses were used to compare ADI-R subscale means due to non-normality.

## RESULTS

35 children with NF1 and 29 TD participants were recruited. **Table 1** provides further details of the demographic characteristics and number of participants per measure. 32 of the participants with NF1 had an inherited pathogenic variant, 2 arose de novo and 1 had an unknown mechanism of inheritance.

### Trajectories of cognitive and adaptive behavioural development

**Table 2** summarises the results of the linear mixed models for each of the measures described below and post-hoc statistics are presented in **Supplementary Materials**.

On the MSEL, the developmental trajectory of children with NF1 differed significantly compared to the trajectory of TD children across all subscales (Visual Reception, Fine Motor, Receptive Language, Expressive Language, Early Learning Composite), with slower progress in the NF1 group. There was also a significant effect of Timepoint within the model; cognitive skills increased for all children as they aged, irrespective of group membership. The interaction between Group and Timepoint also reached statistical significance for all MSEL subscales (**Figure 1**). Pairwise comparisons showed significant differences between the NF and TD groups on all MSEL domains at 5, 24 and 36 months. Only lower Fine Motor skills were observed at 10 months in the NF1 group, with no differences between the groups at 14 months.

**Figure 1:**
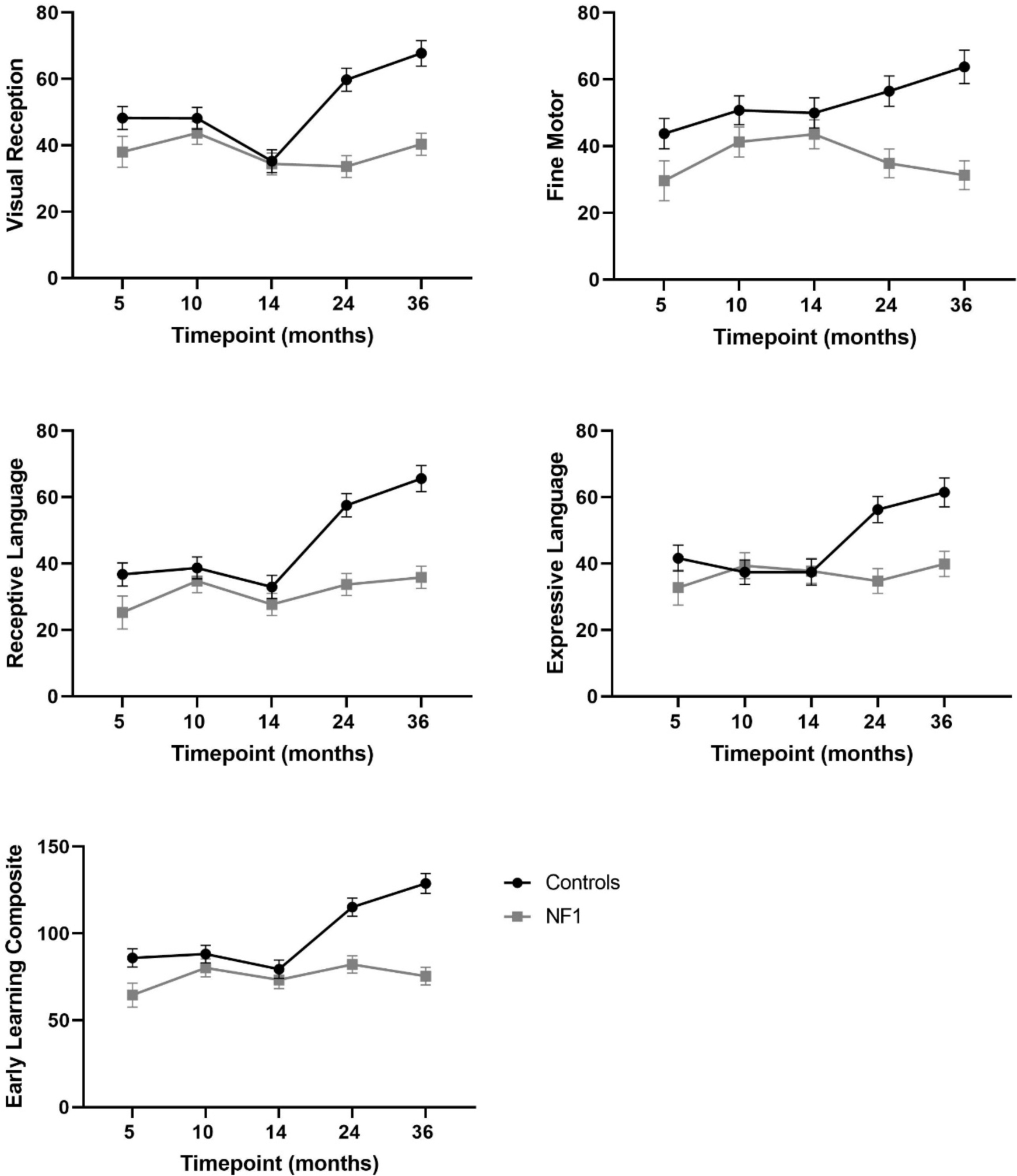
Estimated marginal mean scores of NF1 and TD groups on the Mullen Scale of Early Learning at 5, 10, 14, 24 and 36 months. Error bars represent 95% confidence intervals.

The trajectories of adaptive behavioural skills development were significantly different in NF1 when compared with TD children on four subscales of the VABS; Communication abilities, Daily living skills, Motor Skills and the Adaptive behaviour Composite domain, with slower progress in the NF1 group. However, there was no significant difference between children with NF1 and TD children on the Socialization domain. Adaptive behavioural skills increased significantly for all children over time irrespective of group membership. The interaction between Group and Timepoint also reached statistical significance for all VABS subscales (**Figure 2**). Pairwise comparisons showed that the NF1 participants scored significantly lower than TD participants in Daily living skills, Motor skills and the Adaptive Behavior Composite at 10 months, on all VABS domains at 24 months, but only on communication at 36 months.

**Figure 2:**
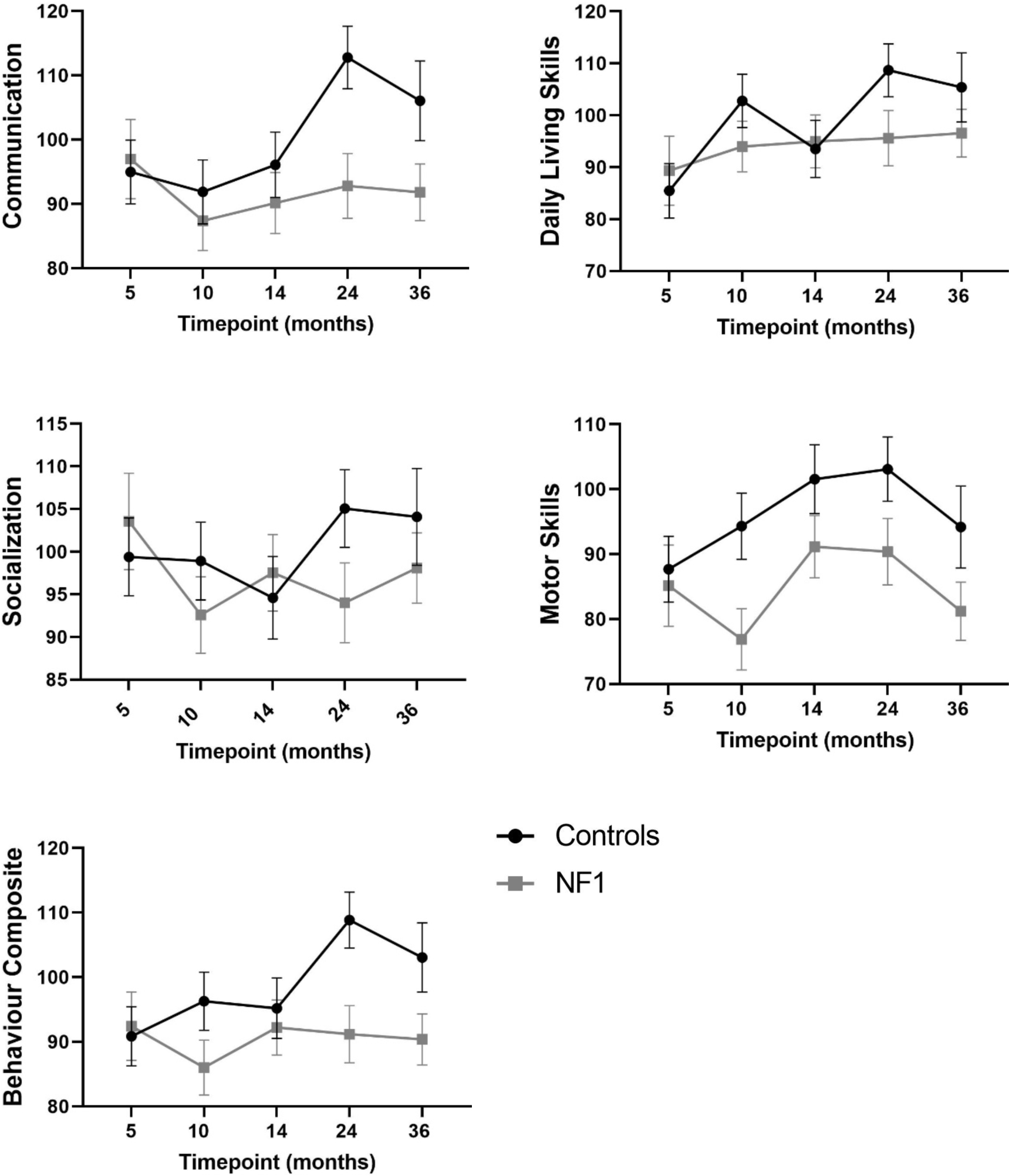
Estimated marginal mean scores of NF1 and TD groups on the Vineland Adaptive Behaviour Scale at 5, 10, 14, 24 and 36 months. Error bars represent 95% confidence intervals.

### ADHD trait development

Children with NF1 showed higher mean levels of ADHD traits on the CBCL DSM-ADHD subscale than TD children **(Figure 3 and Table 2)**. 10.5% of NF1 participants at 36 months had a T-score over 65, suggesting clinically significant ADHD traits. There was no significant effect of Age or interaction between group and Age within this model, as the trajectory in children with NF1 paralleled the trajectory of TD children over time, suggesting similar patterns of change over time. There was no significant difference in CBCL score between male and female participants.

**Figure 3:**
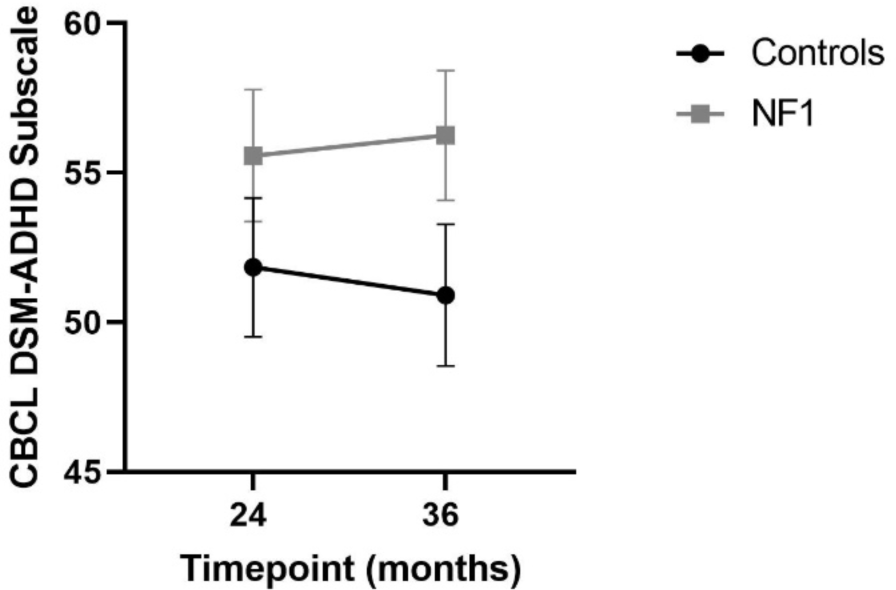
Estimated marginal mean scores of NF1 and TD groups on the CBCL-DSM ADHD Subscale (T scores used) at 24 and 36 months. Error bars represent 95% confidence intervals.

### Autism trait development

Mean total ADOS-2 scores were higher in the NF1 group as compared to the TD group. 48% of NF1 participants at 24 months scored above threshold for autism on the ADOS-2 or BOSA-MV instruments, compared to 0% of TD children (X^2^ (1) = 15.51, p<0.001) (**Table 3**). More male than female participants reached instrumental threshold for autism in the NF1 group (X^2^ (1) = 6.24, p=0.01).

At 36 months, 31% of participants in the NF1 group scored above threshold for autism on the ADOS-2 or BOSA. This was not statistically significant (X^2^ (1) = 0.21, p=0.65), as 25% of TD participants also scored above threshold (although no TD participants gained an eventual research classification of autism) (**Table 3**). There were no statistically significant sex differences between the autism and non-autism participants in either the NF1 (X^2^ (1) = 2.52, p=0.11) or TD groups (X^2^ (1) = 1.11, p=0.29).

On the ADI-R at 36 months, there were significant differences, with higher mean scores on each of the 4 subscales in NF1 compared to TD participants (p<0.001) (**Figure 4 and Table 3**).

**Figure 4:**
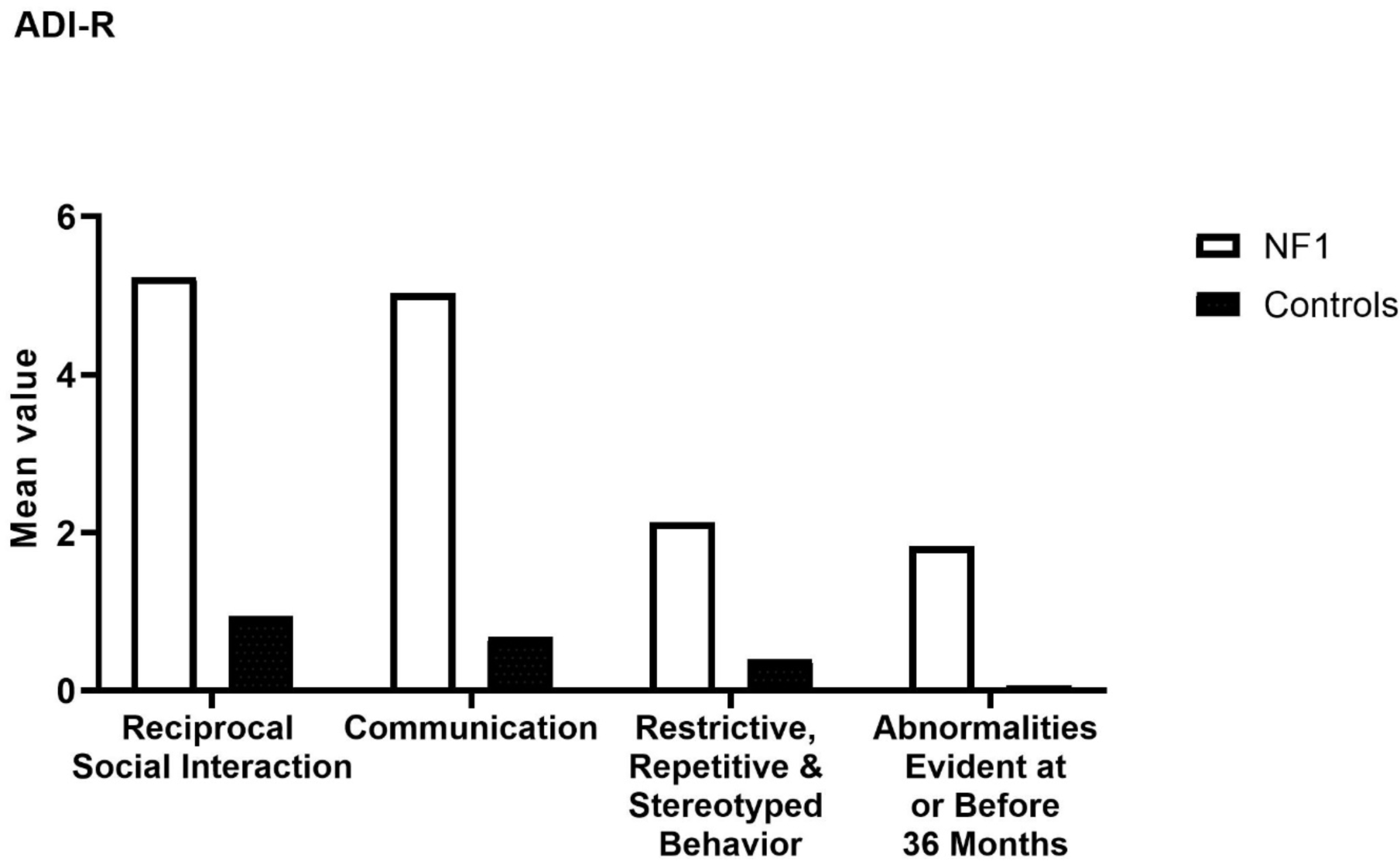
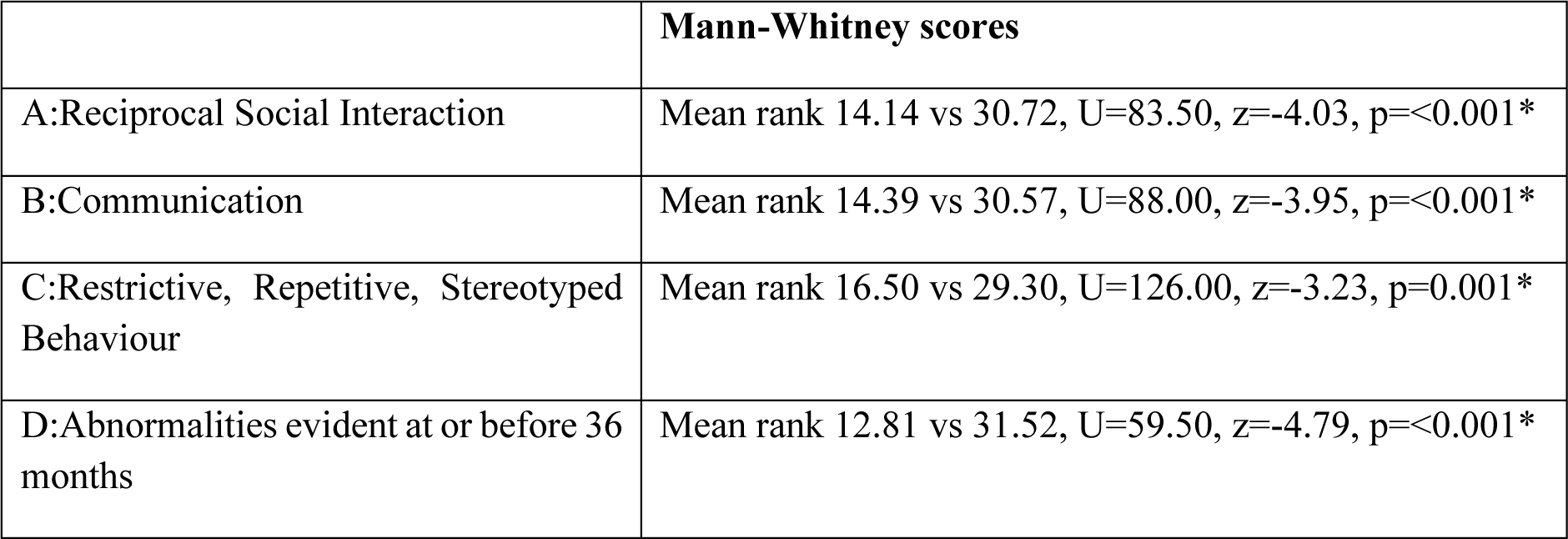
Mean values and Mann-Whitney results for subscales A-D on the Autism Diagnostic Interview-Revised (ADI-R). A – qualitative abnormalities in reciprocal social interaction, B – qualitative abnormalities in communication, C – restricted, repetitive and stereotyped patterns of behaviour and D – abnormalities of development evident at or before 36 months.

When combining the ADI-R and ADOS-2/BOSA at 36 months to give a research classification of autism, as detailed in the **Supplementary materials**, 14% of the NF1 cohort met criteria for autism compared to 0% of TD participants; this was also statistically not significant (X^2^ (1) = 2.71, p=0.10) (**Table 3**).

## DISCUSSION

To our knowledge, this is the first study to systematically investigate the developmental trajectories of cognitive and behavioural development in children with NF1 from 5 to 36 months of age using both objective and parental report measures. Our study suggests significantly lower trajectories of cognitive, motor, language and adaptive behaviour development in the NF1 group, with an overall mean difference that strengthens over developmental time and is clear by 24 months of age.

On the MSEL, lower scores are first evident at 5 months, but these differences are strengthened by 24 months, and remain at 36 months. These results are consistent with a previous longitudinal study which showed significantly lower cognitive function in NF1 at 21 months of age, which remained at the follow-up evaluation at 40 months.^14^ It is interesting to note that only subtle group differences in cognitive functioning were observed at 10 and 14 months. Our previous reports on the same cohort suggest differences arising in infancy in early auditory habituation and visual attention. Using an auditory habituation paradigm, Begum-Ali et al (2021) found developmental differences in auditory processing in infants with NF1 at 10 and 14 months, possibly suggestive of alterations in early sensory processing and specialisation.^16^ Such early-stage processing differences that typically present in the first year of life may represent the beginning of a series of compensatory and adaptive brain processes that trigger an alternative trajectory of subsequent development.^25,26^ Further, based on the results of the parent rated VABS, there may be differences in the pace of developmental change in the NF1 group with periods of ‘catch-up’ over time.

Our results indicate that levels of ADHD traits are higher in the NF1 group but were in the clinically significant range for only 10.5% at 36 months. Behavioural features of ADHD generally tend to peak in the school-age period, which has also been noted in NF1 with rates of ADHD as high as 50%.^6^ Animal model studies suggest that dopaminergic dysfunction in NF1 underlies attentional deficits.^27^ Clinically, the ADHD symptomatology seen in NF1 is very similar to idiopathic ADHD. However, in-depth analyses of neurophysiological processes underlying attention difficulties in NF1 suggests differences in cognitive control processes. Further understanding of these processes will be needed to move away from generic pharmacological intervention for ADHD and develop more personalised approaches. Of note, only 3 participants at 36 months gained a T-score >65 however all of these participants also met research criteria for autism, in keeping with previous research indicating that autism and ADHD often co-occur in NF1 children.^6^

We hypothesised that higher proportions of the NF1 cohort would demonstrate autism traits on administered instruments at 24 and 36 months compared to TD participants. Almost half of the NF1 sample met instrumental threshold for autism at 24 months on the observer rated measures, but this was somewhat attenuated at 36 months (31%). When combining both the parental interview (ADI-R) and the observer rated measures (ADOS-2 or BOSA-MV), 14% of the NF1 sample met research classification for autism at 36 months. These results were statistically non-significant (most likely due to sample size), however they are consistent with previous studies which suggested rates of co-occurring autism in NF1 between 10-25%.^7^ Given that the autism behavioural phenotype in NF1 is broadly similar to idiopathic autism,^28^ it will be important to identify similarities and differences in early-stage markers in the two cohorts.

Consistent with a previous longitudinal study of NF1 children aged 21-40 months,^13^ we largely found no sex differences in the trajectories of cognitive, behavioural or social development. These sex differences may emerge at a later timepoint, as there are significant sex differences observed in cross-sectional studies in school-age children with NF1. Males are more likely to demonstrate differences in learning and social functioning, mirroring the pattern seen in the general population.^5,29^ Longitudinal studies of infants with high familial likelihood of neurodevelopmental conditions such as autism or ADHD have found that females in general perform better than males in all dimensions of cognitive functioning.^30^ Alternatively, sex differences may not have been noted in our sample due to the relatively small sample size.

Methodological strengths of our study included tracking of a prospectively ascertained sample of children with NF1. Limitations include a relatively small sample size due to variable retention and rolling recruitment. The NF1 group was composed primarily of children with an inherited pathogenic variant, due to the early age at which assessments began. Our TD group showed a higher than expected proportion of participants reaching threshold for autism at 36 months on the ADOS-2/BOSA, however when combined with the ADI-R, none of the TD group was given a research classification of autism. Due to COVID limitations we utilised the BOSA for some participants, which is administered online and may not gather as accurate a picture of traits as ADOS-2. Finally, research thresholds using the ADI-R and ADOS/BOSA rather than gold-standard clinical best estimates were used for the autism classification in the NF1 cohort.

## CONCLUSION

Our results collectively suggest that the NF1 brain development is atypical, with early-stage sensory processing difficulties seen in infancy consolidated to behavioural phenotypic differences by 24 months.^15,16^ Clinically, this highlights that developmental monitoring and referral for early interventions should be considered by the age of 2 in children with NF1. Intervention targeting neurocognitive modifiers such as executive attention or social engagement may ameliorate the impact of genetic or environmental vulnerabilities on the developing brain.^29^ Future work should include replication of our findings in larger cohorts, investigating the similarities and differences in the developmental profiles seen in NF1 to other cohorts of infants at higher likelihood of common neurodevelopmental conditions such as autism and ADHD. This will inform potential intervention development.

## Data Availability

Deidentified participant data is available through the STAARS network via data sharing procedures that comply with ethical requirements, due to the sensitive nature of the data collected. Available at: https://www.basisnetwork.org/. There is no known end date of data availability.

## Abbreviations

NF1: neurofibromatosis type 1
ADHD: attention deficit hyperactivity disorder
MSEL: Mullen Scales of Early Learning
VABS: Vineland Adaptive Behavior Scales
ADOS-2: Autism Diagnostic Observation Schedule
BOSA-MV: Brief Observation of Symptoms of Autism for Minimally Verbal children
ADI-R: Autism Diagnostic Interview-Revised
CBCL: Child Behavior Checklist.

## Acknowledgments

We are extremely grateful to the families who have given their time for our research, and NF charities particularly Nerve Tumours UK and Childhood Tumour Trust. We would like to thank the researchers who assisted with recruitment and data collection; Kim Davies, Janice Fernandes, Marian Greensmith, Natalie Vaz, Claire Bennett, Olivia Mitchell, Imogen Crook, Amelia Pearson, Henna Ahmed and Sofia Ahmed. We are grateful to the placement students who assisted with data collection; Francesca Conti, Zoë Freeman and Meg Jackson.

## Members of the EDEN-STAARS team

The STAARS team includes: Mary Agyapong, Tessel Bazelmans, Leila Dafner, Mutluhan Ersoy, Teodora Gliga, Amy Goodwin, Rianne Haartsen, Hanna Halkola, Alexandra Hendry, Rebecca Holman, Sarah Kalwarowsky, Sarah Lloyd-Fox, Luke Mason, Nisha Narvekar, Laura Pirazzoli and Chloë Taylor.

The EDEN team includes: Grace Vassallo, Judith Eelloo, D. Gareth Evans, Siobhan West, Eileen Hupton, Louise Robinson, Neeta Lakhani, Brian Wilson, Deborah Osio, Charles Shaw-Smith, Natalie Canham and Saba Sharif.

## Supplementary materials

### Procedure

The study assessments took place at the Division of Psychology and Mental Health, University of Manchester, and the Centre for Brain and Cognitive Development, Birkbeck, University of London. Prior written informed consent was obtained from the parent. Testing took place if the child was physically well and content. Participant families were provided with reimbursement for expenses. The behavioural measures described below were part of a more comprehensive experimental protocol.

### Measures

*ADOS-2*: Based on the expressive language ability of the participants, either the Toddler module, Module 1 or Module 2 of the ADOS-2 were used at 24 and 36 months (**Table 2**). Coding was carried out from videos, with an inter-rater reliability of 79.1% for the NF1 cohort.

*BOSA-MV*: Based on the expressive language ability of the participants, either the Toddler module or Module 1 of the BOSA-MV were used at 24 and 36 months (**Table 2**).

*ADI-R*: Four subscales are produced: A – qualitative abnormalities in reciprocal social interaction, B – qualitative abnormalities in communication, C – restricted, repetitive and stereotyped patterns of behaviour and D – abnormalities of development evident at or before 36 months. Each subscale has an algorithm score cut-off for Autism.

### Classification of autism

In our paper, the following thresholds are used on the ADOS-2 and BOSA-MV to indicate autism traits at 24 months:

- ‘mild-to-moderate’ or ‘moderate-to-severe’ concern on the ADOS-2 Toddler module
- ‘autism-spectrum’ or ‘autism’ on ADOS-2 Module 1
- a score of 6 on the BOSA-MV toddler module or a score of 5 on the BOSA-MV Module 1.

Luyster et al (2009) state that at least 95% of children with Autism Spectrum Disorder and no more than 10% of typically developing children would fall into the two groups suggesting clinical concern on the ADOS Toddler module (mild-to-moderate and moderate-to-severe).^22^ Dow et al (2022) recommend a cut-off of 6 for Autism Spectrum Disorder for the BOSA-MV toddler module and a score of 5 as a cut-off for the BOSA-MV Module 1.^23^

#### Participants were assigned a research instrumental classification of autism at 36 months if they scored either

- ‘autism-spectrum’ or ‘autism’ on ADOS-2 Module 1 or 2
- a cut off of 5 on the BOSA-MV Module 1

AND

- Met threshold for subscale A and came within one point of B, or met B and came within one point of A on the ADI-R, as suggested by Risi et al (2006).^31^

### Study numbers

Flowchart showing the number of NF1 and TD participants at each time point in the study. Flexible recruitment and variable retention across visits meant that the sample size varied between time points.

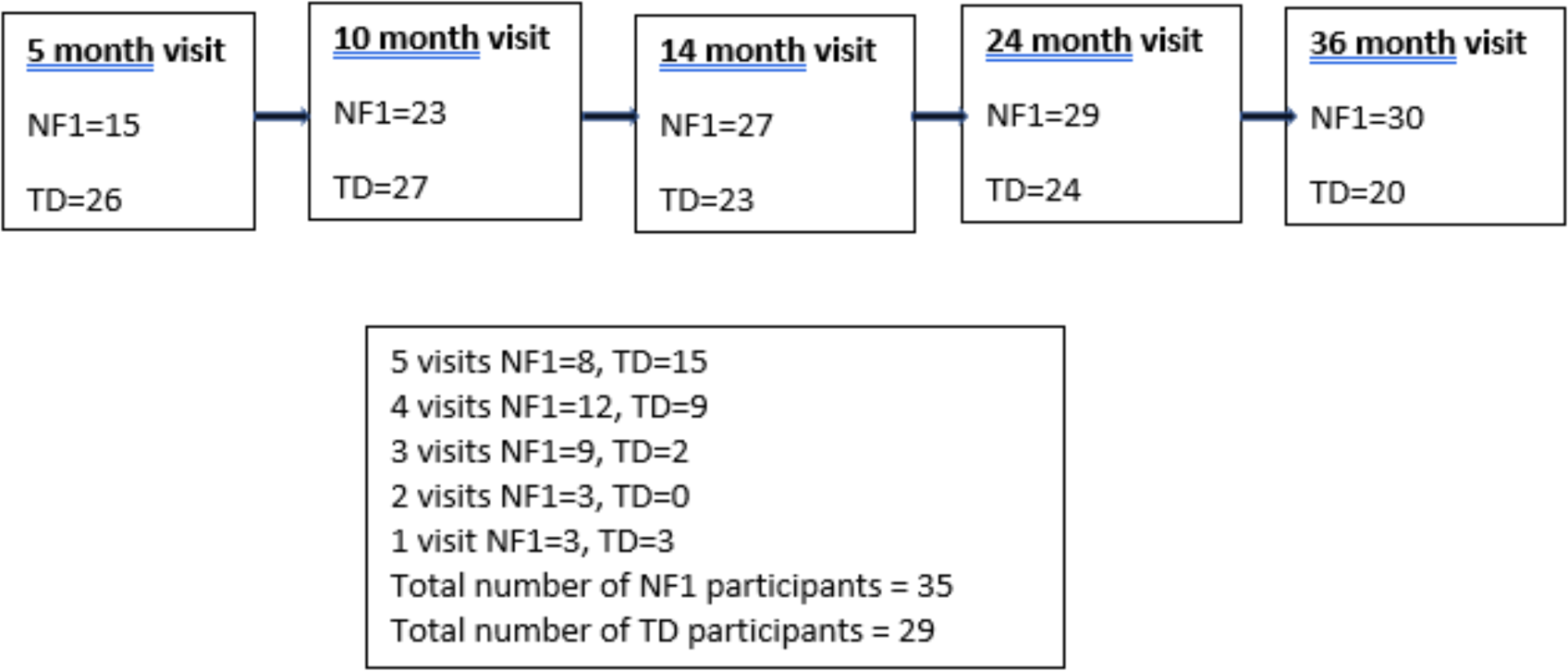

### Descriptive statistics including means and SDs for the NF1 and TD group at the five assessment time points for Mullen Scales of Early Learning and Vineland Adaptive Behaviour Scale

|  | 5 Months |  |  |  | 10 Months |  |  |  | 14 Months |  |  |  | 24 months |  |  |  | 36 months |  |  |  |
| --- | --- | --- | --- | --- | --- | --- | --- | --- | --- | --- | --- | --- | --- | --- | --- | --- | --- | --- | --- | --- |
|  | TD group |  | NF1 Group |  | TD group |  | NF1 Group |  | TD group |  | NF1 Group |  | TD group |  | NF1 Group |  | TD group |  | NF1 Group |  |
|  | *n* | *Mean*<br>*[SD]* | *n* | *Mean*<br>*[SD]* | *n* | *Mean*<br>*[SD]* | *n* | *Mean*<br>*[SD]* | *n* | *Mean*<br>*[SD]* | *n* | *Mean*<br>*[SD]* | *n* | *Mean*<br>*[SD]* | *n* | *Mean*<br>*[SD]* | *n* | *Mean*<br>*[SD]* | *n* | *Mean*<br>*[SD]* |
| **Mullen Scale of Early Learning T scores** |  |  |  |  |  |  |  |  |  |  |  |  |  |  |  |  |  |  |  |  |
| Gross Motor | 26 | 43.69 [9.41] | 15 | 37.40 [9.72] | 27 | 34.89 [11.78] | 23 | 30.00 [10.25] | 23 | 36.74 [13.38] | 27 | 31.96 [12.75] | - | - | 26 | 35.04 [8.45] | - | - | - | - |
| Visual reception | 26 | 47.27 [6.33] | 15 | 37.47 [11.53] | 27 | 48.85 [7.99] | 23 | 43.96 [8.08] | 23 | 35.09 [8.88] | 27 | 33.67 [5.17] | 24 | 59.63 [10.66] | 27 | 33.11 [9.83] | 19 | 67.58 [9.34] | 27 | 40.63 [8.65] |
| Fine motor | 26 | 42.92 [10.46] | 15 | 32.8 [6.99] | 27 | 51.63 [12.88] | 23 | 42.13 [11.29] | 23 | 49.65 [12.18] | 27 | 43.85 [11.07] | 24 | 56.00 [12.98] | 27 | 33.89 [10.49] | 19 | 64.68 [11.32] | 27 | 31.22 [9.67] |
| Receptive language | 26 | 36.88 [11.28] | 14 | 25.71 [9.31] | 27 | 39.26 [8.96] | 23 | 34.87 [7.73] | 23 | 32.87 [6.50] | 27 | 28.33 [6.01] | 24 | 57.67 [8.73] | 27 | 33.85 [10.65] | 19 | 65.84 [6.30] | 27 | 35.89 [9.70] |
| Expressive language | 26 | 41.85 [7.45] | 15 | 35.13 [6.35] | 27 | 36.85 [9.89] | 23 | 39.35 [12.56] | 23 | 37.09 [8.81] | 27 | 38.19 [10.18] | 24 | 55.42 [12.30] | 27 | 34.52 [8.70] | 19 | 60.84 [8.51] | 26 | 39.50 [10.57] |
| Early Learning Composite Scores | 26 | 85.00[9.32] | 14 | 67.86 [9.46] | 27 | 88.89 [12.19] | 23 | 81.13 [10.70] | 23 | 78.78 [11.99] | 27 | 73.67 [7.30] | 24 | 114.25[17.91] | 27 | 81.48[17.91] | 19 | 129.05[11.75] | 26 | 75.88[13.45] |

| **Vineland Adaptive Behaviour Scale Standard Scores** |  |  |  |  |  |  |  |  |  |  |  |  |  |  |  |  |  |  |  |  |
| --- | --- | --- | --- | --- | --- | --- | --- | --- | --- | --- | --- | --- | --- | --- | --- | --- | --- | --- | --- | --- |
| Communication |  |  |  |  |  |  |  |  |  |  |  |  |  |  |  |  |  |  |  |  |
| Daily living skills | 22 | 94.95 [10.91] | 14 | 95.71 [14.04] | 21 | 92.86 [12.76] | 23 | 86.13 [14.91] | 19 | 96.11 [10.49] | 24 | 91.04 [10.08] | 22 | 112.14 [8.82] | 22 | 93.41 [7.5] | 12 | 105.08 [8.58] | 30 | 90.73 [14.23] |
| Socialisation | 22 | 86.32 [19.86] | 14 | 92.71 [18.26] | 22 | 102.45 [10.76] | 23 | 93.74 [9.14] | 18 | 93.44 [8.39] | 23 | 95.52 [10.85] | 22 | 108.36 [7.36] | 22 | 96.14 [9.25] | 12 | 104 [10.06] | 30 | 97.9 [14.67] |
| Motor | 23 | 98.70 [12.26] | 14 | 101.36 [14.21] | 22 | 98.77 [10.04] | 21 | 92.76 [14.59] | 18 | 94.22 [9.79] | 23 | 97.96 [9.43] | 22 | 104.64 [7.86] | 22 | 94.55 [7.71] | 12 | 103.25 [6.05] | 30 | 98.47 [15.78] |
| Adaptive behaviour composite | 21 | 88.67 [12.99] | 14 | 85.07 [13.37] | 21 | 95.76 [16.86] | 23 | 79.91 [14.86] | 18 | 101.67 [9.49] | 24 | 93.42 [11.14] | 22 | 102.41 [7.93] | 22 | 93.05 [8.67] | 12 | 92 [8.52] | 30 | 83.77 [13.13] |
|  | 19 | 90.63 [11.93] | 14 | 92.29 [11.13] | 20 | 96.80 [10.44] | 21 | 85.86 [12.85] | 17 | 95.29 [9.73] | 23 | 93.17 [9.36] | 22 | 108.05 [8.16] | 22 | 92.73 [5.78] | 12 | 100.92 [8.58] | 30 | 91.4 [14.62] |

\*Missing data not included in n, mean or SD but kept in model for imputation

### Post-hoc T-test analysis for the NF1 and TD group means at the five assessment time points for Mullen Scales of Early Learning and Vineland Adaptive Behaviour Scale. Bonferroni corrected alpha value =<0.01

|  | 5 Months T-test<br>(NF1 group – TD group) | 10 Months T-test<br>(NF1 group – TD group) | 14 Months T-test<br>(NF1 group – TD group) | 24 Months T-test<br>(NF1 group – TD group) | 36 Months T-test<br>(NF1 group – TD group) |
| --- | --- | --- | --- | --- | --- |
| **Mullen Scale of Early Learning T Scores** |  |  |  |  |  |
| Visual Reception | t(39)=-3.53, p<0.001* | t(48)=-2.15, p=0.037 | t(48)=-0.70, p=0.485 | t(49)=-9.24, p<0.001* | t(44)=-10.07, p<0.001* |
| Fine Motor | t(39)=-3.34, p=0.002* | t(48)=-2.75, p=0.008* | t(48)=-1.76, p=0.084 | t(49)=-6.72, p<0.001* | t(44)=-10.77, p<0.001* |
| Receptive Language | t(38)=-3.17, p=0.003* | t(48)=-1.84, p=0.072 | t(48)=-2.56, p=0.014 | t(49)=-8.67, p<0.001* | t(44)=-11.80, p<0.001* |
| Expressive Language | t(39)=-2.93, p=0.006* | t(48)=0.79, p=0.436 | t(48)=0.40, p=0.688 | t(49)=-7.06, p<0.001* | t(44)=-7.24, p<0.001* |
| Early Learning Composite Scores | t(38)=-5.52, p<0.001* | t(48)=-2.37, p=0.022 | t(48)=-1.85, p=0.07 | t(49)=-6.41, p<0.001* | t(44)=-13.80, p<0.001* |
| **Vineland Adaptive Behaviour Scale Standard Scores** |  |  |  |  |  |
| Communication | t(34)=0.182, p=0.857 | t(42)=-1.601, p=0.117 | t(41)=0.79, p=0.116 | t(42)=-7.59, p<0.001* | t(40)=-3.25, p<0.001* |
| Daily Living Skills | t(34)=0.971, p=0.338 | t(43)=-2.933, p=0.005* | t(39)=0.67, p=0.507 | t(42)=-4.85, p<0.001* | t(40)=-1.32, p=0.195 |
| Socialization | t(35)=0.603, p=0.550 | t(41)=-1.580, p=0.122 | t(39)=1.237, p=0.223 | t(42)=-4.30, p<0.001* | t(40)=-1.02, p=0.316 |
| Motor Skills | t(33)=-0.793, p=0.433 | t(42)=0.585, p<0.001* | t(40)=-2.527, p=0.16 | t(42)=-3.74, p<0.001* | t(40)=-2.00, p=0.052 |
| Adaptive Behaviour Composite | t(31)=0.405, p=0.688 | t(39)=-2.983, p<0.005* | t(38)=-0.696, p=0.490 | t(42)=-7.19, p<0.001* | t(40)=-2.11, p=0.042 |

#### MSEL post hoc tests

**NF1 vs TD at 5 months:** Visual Reception [t(39)=-3.53, p<0.001, CI (-15.42 to -4.18)], Fine Motor [t(39)=-3.34, p=0.002, CI (-16.26 to -3.98)], Receptive Language [t(38)=-3.17, p=0.003, CI (-18.31 to -4.03)], Expressive Language [t(39)=-2.93, p=0.006, CI (-11.35 to -2.07)], Early Learning Composite [t(38)=-5.52, p<0.001, CI (-23.43 to -10.86)]. **NF1 vs TD at 10 months:** Visual Reception [t(48)=-2.15, p=0.037, CI (-9.48 to -0.31)], Fine Motor [t(48)=-2.75, p=0.008, CI (-16.45 to -2.55)], Receptive Language [t(48)=-1.84, p=0.072, CI (-9.19 to 0.41)], Expressive Language [t(48)= 0.79, p=0.436, CI (-3.89 to 8.88)], Early Learning Composite [t(48)=-2.37, p=0.022, CI (-14.34 to -1.18)]). **NF1 vs TD at 14 months:** Visual Reception [t(48)=-0.70, p=0.485, CI (-5.48 to 2.64)], Fine Motor [t(48)=-1.76, p=0.084, CI (-12.41 to 0.81)], (Receptive Language [t(48)=-2.56, p=0.014, CI (-8.09 to -0.98)], Expressive Language [t(48)=0.40, p=0.688, CI (-4.36 to 6.56)], Early Learning Composite [t(48)=-1.85, p=0.07, CI (-10.67 to 0.44)]. **NF1 vs TD at 24 months:** Visual Reception [t (49)=-9.24, p<0.001, CI (-32.28 to -20.75)], Fine Motor [t(49)=-6.72, p <0.001, CI (-28.72 to -15.50)], Receptive Language [t(49)=-8.67, p <0.001, CI (-29.34 to -18.29)], Expressive Language [t(49)=-7.06, p <0.001, CI (-26.84 to -14.95)], Early Learning Composite [t(49)=-6.41, p <0.001, CI (-43.04 to -22.50)]. **NF1 vs TD at 36 months:** Visual Reception [t(44) =-10.07, p <0.001, CI (-32.35 to -21.55)], Fine Motor [t(44)=-10.77, p <0.001, CI (-39.73 to -27.20)], Receptive Language [t(44)=-11.80, p <0.001, CI (-35.07 to -24.84)], Expressive Language [t(44)=-7.24, p <0.001, CI (-27.29 to -15.40)], Early Learning Composite [t(44)=-13.80, p <0.001, CI (-60.94 to -45.40)].

#### VABS post hoc tests

**NF1 vs TD at 5 months:** Communication [t(34)=0.182, p=0.857, CI (-7.72 to 9.24)], Daily living skills [t(34)=0.971, p=0.338, CI (-6.99 to 19.78)], Socialization [t(35)=0.603, p=0.550, CI (-6.30 to 11.62)], Motor Skills [t(33)=-0.793, p=0.433, CI (-12.82 to 5.63)] and Adaptive behaviour Composite [t(31)=0.405, p=0.688, CI (-6.68 to 9.99)]. **NF1 vs TD at 10 months:** Communication [t(34)=0.182, p=0.857, CI (-15.2 to 1.80)], Daily living skills [t(43)=-2.933, p=0.005, CI (-14.7 to -2.72)], Socialization [t(35)=0.603, p=0.550, CI (-13.69 to 1.67)], Motor Skills [t(42)=0.585, p<0.001, CI (-28.5 to -9.2)] and Adaptive behaviour Composite [t(39)=-2.983, p<0.005, CI (-18.36 to -3.52)]. **NF1 vs TD at 14 months:** Communication [t(41)=0.79, p=0.116, CI (-11.43 to 1.3)], Daily living skills [t(39)=0.67, p=0.507, CI (-4.20 to 8.35)], Socialization [t(39)=1.237, p=0.223, CI (-2.37 to 9.84)], Motor Skills [t(40)=-2.527, p=0.16, CI (-14.85 to -1.65)] and Adaptive behaviour Composite [t(38)=-0.696, p=0.490, CI (-8.28 to 4.04)]. **NF1 vs TD at 24 months:** Communication [t(42)=-7.59, p <0.001, CI (-23.71 to -13.75)], Daily living skills [t(42)=-4.85, p <0.001, CI (-17.31 to -7.14)], Socialization [t(42)=-4.30, p <0.001 CI (-14.83 to -5.36)], Motor Skills [t(42)=-3.74, p <0.001, CI (-14.42 to -4.31)] and Adaptive behaviour Composite [t(42)=-7.19, p <0.001, CI (-19.62 to -11.02)]. **NF1 vs TD at 36 months:** Communication [t(40)=-3.25, p <0.001, CI (-23.27 to -5.43)], Daily living skills [t(40)=- 1.32, p 0.195, CI (-15.46 to 3.26)], Socialization [t(40)=-1.02, p 0.316, CI (-14.31 to 4.75)], Motor Skills [t(40)=-2.00, p 0.052, CI (-16.54 to 0.08)] and Adaptive behaviour Composite [t(40)=-2.11, p 0.042, CI (-18.65 to -0.38)].

